# Neurokinin-1 receptor antagonist tradipitant improves itch associated with mild atopic dermatitis: Results from EPIONE a randomized clinical trial

**DOI:** 10.1101/2020.06.21.20136911

**Authors:** Sarah E Welsh, Changfu Xiao, Michael A Mohrman, Alyssa R Kaden, Jennifer L Brzezynski, Jingyuan Wang, Sandra P Smieszek, Bartlomiej Przychodzen, Sonja Ständer, Christos Polymeropoulos, Gunther Birznieks, Mihael H Polymeropoulos

## Abstract

**Importance:** Safe oral systemic treatments are needed to treat itch associated with atopic dermatitis (AD).

**Objective:** To examine the efficacy and safety of tradipitant, a neurokinin-1 receptor antagonist, in adults with mild to severe AD.

**Design, Setting, and Participants:** EPIONE was a phase 3, randomized, placebo-controlled, double-blind clinical trial conducted from July 09, 2018 to December 27, 2019 at 74 US centers. Patients were adults 18 years or older with worst itch rated ≥7 on the Worst Itch Numerical Rating Scale (WI-NRS) and ≥ 1% body surface area of AD involvement at screening.

**Interventions:** Patients were randomly assigned (1:1) to twice-daily oral placebo or tradipitant (85 mg) for 8 weeks.

**Main Outcomes and Measures:** The primary endpoint was mean improvement in WI-NRS (baseline to Week 8). Secondary endpoints included disease severity improvement measured by SCORing Atopic Dermatitis (SCORAD) index, the Eczema Area and Severity Index (EASI), and validated Investigator Global Assessment for Atopic Dermatitis (vIGA-AD^™^).

**Results:** 341 patients (mean [SD]: age, 41.8 [15.0] years; sex, 243 [64.8%] female) were randomly assigned to placebo (n = 187) or tradipitant (n = 188). EPIONE did not meet its primary endpoint of reduction in pruritus (LS Mean difference (95% CI), −0.2 (−0.8 to 0.4), P = 0.567). However, robust antipruritic effect was observed in patients with mild lesion severity (rated 1 or 2 by the vIGA-AD at baseline, −1.6 (−2.9 to −0.3), P = 0.015). This result was confirmed by daily diary (−2.09 (−3.31 to −0.87), P = 0.001) and observed after one full day of treatment (−0.61 (−1.21 to −0.01), P = 0.0457). Treatment-emergent adverse events (TEAEs) were reported in 63 of 188 (33.5%) tradipitant patients and 43 of 187 (23.0%) placebo patients. TEAEs with > 2% incidence and twice that of placebo included diarrhea (VLY-686 = 5 (2.7%), PBO = 1 (0.5%)), fatigue (VLY-686 = 5 (2.7%), PBO = 0 (0.0%)), and worsening of AD (VLY-686 = 4 (2.1%), PBO = 1 (0.5%)).

**Conclusions and Relevance:** During 8 weeks of treatment, tradipitant was well-tolerated for all study participants, but did not significantly improve worst itch in the overall study population. However, in patients with mild AD at baseline, tradipitant was observed to have a large and rapid antipruritic effect. These data support the role of neurokinin-1 antagonism and substance P signaling in chronic pruritus related to mild AD. If these findings replicate in the on-going phase 3 study, EPIONE2, tradipitant may represent a promising new oral therapy for these mild AD patients.

**Trial Registration:** Clinicaltrials.gov: NCT03568331, https://clinicaltrials.gov/ct2/show/NCT03568331

**Key Points:** *Question:* Is tradipitant, a selective neurokinin-1 receptor antagonist, efficacious and safe for improving worst itch in patients with mild to severe atopic dermatitis (AD)?

*Findings:* In 375 mild to severe AD patients, tradipitant did not significantly improve worst itch. However, in a subgroup of 79 AD patients with mild lesion severity at baseline, tradipitant significantly improved worst itch and sleep during 8 weeks of treatment.

*Meaning:* These data support the role of neurokinin-1 antagonism and substance P signaling in chronic pruritus related to mild AD, and further suggest tradipitant may represent a new oral systemic option for mild AD patients based on the well-tolerated safety profile and improvement in itch and sleep.

## Introduction

Atopic dermatitis (AD) is a relapsing and remitting disease that affects 4.9% of the US population^1^. AD is characterized by intense pruritus that can lead to scratching and eczematous lesions that vary in extent and severity. Over 60% of AD cases are mild, characterized by slight erythema, induration, and lichenification^2,3^. Moderate to severe cases of AD are characterized by clearly perceptible erythema, induration, lichenification, oozing, cracking, flaking, and bleeding of the skin^4^. Despite the wide range of severity, the American Academy of Dermatology defines pruritus as an essential feature for all clinical diagnoses of AD, including in mild AD, where pruritus can be of high intensity^5,6^. Chronic pruritus, pruritus lasting more than 6 weeks, has been reported by 91% of AD patients^7,8^. Chronic pruritus is especially distressing, as it can disturb sleep and contribute to psychological and social morbidity^9^.

The pathophysiology of AD is driven by a combination of skin barrier dysfunction, neuroinflammation, and immune system dysregulation^10,11^. Normally, the outermost layer of the skin is composed of tightly compacted lipid-protein matrices, which minimizes water loss from the body and prevents pathogen and allergen entry^11,12^. A known loss-of-function mutation in the gene Filaggrin can lead to a disruption in the integrity of these matrices^13^. Scratching induced by pruritus can physically damage the skin barrier and promote further inflammation. Activated immune cells (Th2, Th17, Th22) release a variety of cytokines and chemokines, including IL-4, IL-31, and IL-13, that activate other cells such as antigen presenting cells, dendritic cells, basophils, and mast cells to release pro-inflammatory mediators, and lead to further disturbance of epidermal differentiation and barrier damage. These molecules also mediate pruritus by acting on the sensory neurons in the skin^8^. Activated sensory fibers in the epidermis and upper dermis signal itch and burning/stinging pain, and release neuropeptides. The neuropeptides themselves can cause vasodilation, plasma extravasation, and edema. By attraction of T-cells, the neuropeptides also promote inflammation, resulting in the so-called neuroinflammation. Inflammatory responses and genetic predisposition appear to be dependent on level of lesion severity (Smieszek et al. 2020, under revision)^14,15^. Together these factors likely combine to amplify the inflammatory response and AD presentation, and contribute to the propagation of the “itch-scratch cycle”^16^.

Currently, topical steroids, calcineurin inhibitors, and emollients are the most common treatments for AD^3^. Steroids target the dysregulated immune pathways, and emollients aim to address the skin barrier dysfunction^17^. Antihistamines are often used in combination with other therapies to mediate itch^3^, however as the itch associated with AD is non-histaminergic, antihistamines often have an insufficient effect on relieving pruritus^8^. More recently, direct immunomodulatory agents, such as crisaborole and dupilumab, have been developed for moderate to severe AD. Despite the availability of these therapies, over 40% of AD patients rate dissatisfaction with their current treatment plan and identify pruritus as a persistent feature of their disease, with the reduction of itch as the most important treatment goal^18,19^. Therefore, novel, well-tolerated therapies are needed to treat itch.

Elevated substance P (SP) is found in both serum and lesional skin of patients with AD^8,20,21^. SP, a neuropeptide released from the activation of sensory neurons, preferentially binds to the neurokinin-1 (NK-1) receptor and is a known itch mediator^8,22^. The NK-1 receptor is found on mast cells, keratinocytes, and central and peripheral nerve endings^23^. Activation of the NK-1 receptor by SP leads to intracellular second messenger signaling cascades, which control many cellular processes, including neuro-immune modulation, that may play a key role in the pathogenesis of AD^24^. Increases in SP are known to induce secretion of pro-inflammatory mediators, such as TNF-α, IFN-γ, and IL-2^25^. SP signaling in the skin also upregulates the expression of nerve growth factor (NGF), which affects pruriception in the skin^26^. These factors may combine to neuronal hypersensitivity of sensory small fibers towards itch by reducing the threshold of cutaneous nerve fibers for pruritogens. A typical clinical sign for neuronal hypersensitivity is alloknesis (patients scratch and as a result worsen their itching). Preclinical models confirm that administration of NK-1 receptor antagonist decreased scratching behavior in mice^27^. This evidence suggests that inhibition of this signaling pathway via an NK-1 receptor antagonist may lead to reduction of SP-induced pruritus in AD. Tradipitant (VLY-686), a novel NK-1 receptor antagonist, has the potential to reduce itch related to AD through inhibition of SP-mediated itch signaling.

A previous phase 2 study (Study 2102) was conducted at 28 centers in the US^28^. Study 2102 demonstrated that eight weeks of tradipitant treatment improved worst itch and disease severity. These results led to the design of the phase 3 study, EPIONE, studying tradipitant treatment in 375 patients with chronic pruritus associated with AD. The results of EPIONE are presented here as an 8-week, placebo-controlled study to determine the efficacy and safety of twice-daily tradipitant administration in reduction of chronic pruritus in patients with AD.

## Methods

### Study design

EPIONE was a randomized, double-blind, placebo-controlled phase 3 study and was approved by human research ethics committees at all participating institutions. Participants provided written informed consent and were provided a copy of the signed consent form before any screening procedures occurred. The study was conducted across 74 sites in the United States, from July 09, 2018 to December 27, 2019. Participants were randomly assigned to tradipitant (85 mg) or matched placebo, which was taken orally twice-daily (morning and evening) for 8 weeks. No participants (investigators, study staff, patients) knew the treatment assignment, and blinding was maintained throughout the study through database lock.

This study had two phases: the screening phase and the evaluation phase. The screening phase consisted of an initial visit to evaluate eligibility, followed by a wash-out period lasting 14 to 45 days. Participants completed a daily diary throughout this washout period. Participants meeting all eligibility criteria were randomized at the baseline visit and were dispensed study medication. Participants were randomized (1:1) using an automated interactive web response system. Participants who did not meet eligibility criteria were considered screen failures. During the evaluation phase, patients returned to the clinic for four additional study visits where safety and efficacy assessments were performed.

### Participants

Participants were men and women aged between 18 and 70 years, with chronic pruritus related to AD that was refractory to treatment by patient history, pruritus intensity rated by the Worst Itch Numerical Rating Scale (WI-NRS) diary average score of 7 or greater, and ≥ 1% body surface area of AD involvement.

### Assessments and Outcomes

Itch was assessed by the patient by WI-NRS during the study visits. Responses were measured by an 11-point numeric scale with 0 being “No Itching” and 10 being “Worst Itch Imaginable”. Disease severity was assessed by the investigators utilizing the SCORing Atopic Dermatitis (SCORAD) index^29^, the Eczema Area and Severity Index (EASI)^30^, and the validated Investigator Global Assessment for Atopic Dermatitis (vIGA-AD™). Sleep disturbance was measured by daily diary 11-point numerical rating scale (NRS) with 0 being “No Sleep Disruption” and 10 being “Very Severe Sleep Disruption”, 100 mm SCORAD visual analogue scale (VAS), and the Patient-Oriented Eczema Measure (POEM)^31^.

The primary objective was to evaluate the efficacy of tradipitant in reducing worst itch as measured by WI-NRS at Week 8. Secondary objectives included evaluating the response rate of ≥ 4 points on WI-NRS, improvement in disease severity by vIGA-AD, SCORAD, and EASI, improvement in sleep, and to explore the safety and efficacy of multiple oral doses of tradipitant.

Safety assessments included the regular monitoring and recording of all adverse events (AEs) and serious adverse events (SAEs); monitoring of hematology, serum chemistry, and urinalysis values, vital signs, body measurements, and suicidal ideation and behavior; and the performance of physical examinations and ECGs.

### Statistical analyses

375 participants were enrolled in EPIONE. This sample size was determined based on previous historical data. Statistical analyses were performed using two-sided tests; significance was considered at P ≤ 0.05.

The intent-to-treat (ITT) population was defined as all individuals randomized into the study who received a dose of study drug and completed at least one post-baseline efficacy measurement while on study medication. The primary analysis for efficacy used mixed model repeated measures (MMRM) analysis. The responder analysis was based on a two-sided Fisher’s Exact Test. All analyses and tabulations were performed using SAS®.

## Results

EPIONE screened 1015 individuals and enrolled 375 patients meeting the inclusion criteria, with 286 patients completing the study (**Figure 1**). The ITT population consisted of 341 patients (VLY-686 = 171, PBO = 170). There were no significant differences across treatment groups in baseline demographics and disease severity (**Table I**).

**Table I.**
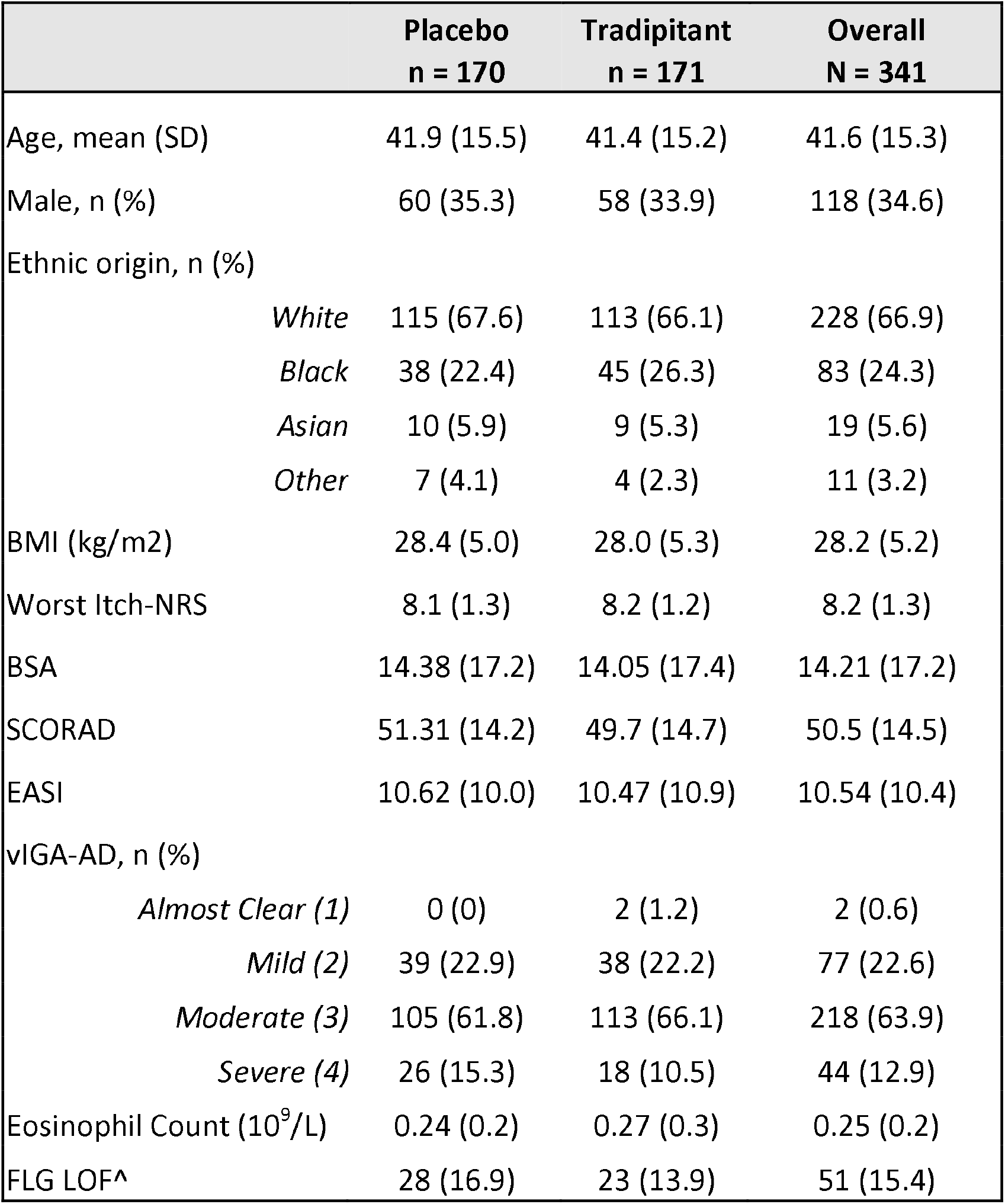
Baseline Demographic and Clinical Characteristics. Data are in mean (SD) or number (%). BMI=body mass index, NRS=numerical rating scale, SCORAD=SCORing Atopic Dermatitis index, EASI=Eczema Area and Severity Index, vIGA-AD=validated Investigator Global Assessment for Atopic Dermatitis, BSA=body surface area ^VLY-686 n = 166, PBO n = 166, Overall N = 322

|  | Placebo<br>n = 170 | Tradipitant<br>n = 171 | Overall<br>N = 341 |
| --- | --- | --- | --- |
| Age, mean (SD) | 41.9 (15.5) | 41.4 (15.2) | 41.6 (15.3) |
| Male, n (%) | 60 (35.3) | 58 (33.9) | 118 (34.6) |
| Ethnic origin, n (%) |  |  |  |
| *White* | 115 (67.6) | 113 (66.1) | 228 (66.9) |
| *Black* | 38 (22.4) | 45 (26.3) | 83 (24.3) |
| *Asian* | 10 (5.9) | 9 (5.3) | 19 (5.6) |
| *Other* | 7 (4.1) | 4 (2.3) | 11 (3.2) |
| BMI (kg/m<sup>2</sup>) | 28.4 (5.0) | 28.0 (5.3) | 28.2 (5.2) |
| Worst Itch-NRS | 8.1 (1.3) | 8.2 (1.2) | 8.2 (1.3) |
| BSA | 14.38 (17.2) | 14.05 (17.4) | 14.21 (17.2) |
| SCORAD | 51.31 (14.2) | 49.7 (14.7) | 50.5 (14.5) |
| EASI | 10.62 (10.0) | 10.47 (10.9) | 10.54 (10.4) |
| vIGA-AD, n (%) |  |  |  |
| *Almost Clear (1)* | 0 (0) | 2 (1.2) | 2 (0.6) |
| *Mild (2)* | 39 (22.9) | 38 (22.2) | 77 (22.6) |
| *Moderate (3)* | 105 (61.8) | 113 (66.1) | 218 (63.9) |
| *Severe (4)* | 26 (15.3) | 18 (10.5) | 44 (12.9) |
| Eosinophil Count (10<sup>9</sup>/L) | 0.24 (0.2) | 0.27 (0.3) | 0.25 (0.2) |
| FLG LOF<sup>^</sup> | 28 (16.9) | 23 (13.9) | 51 (15.4) |

**Figure 1.**
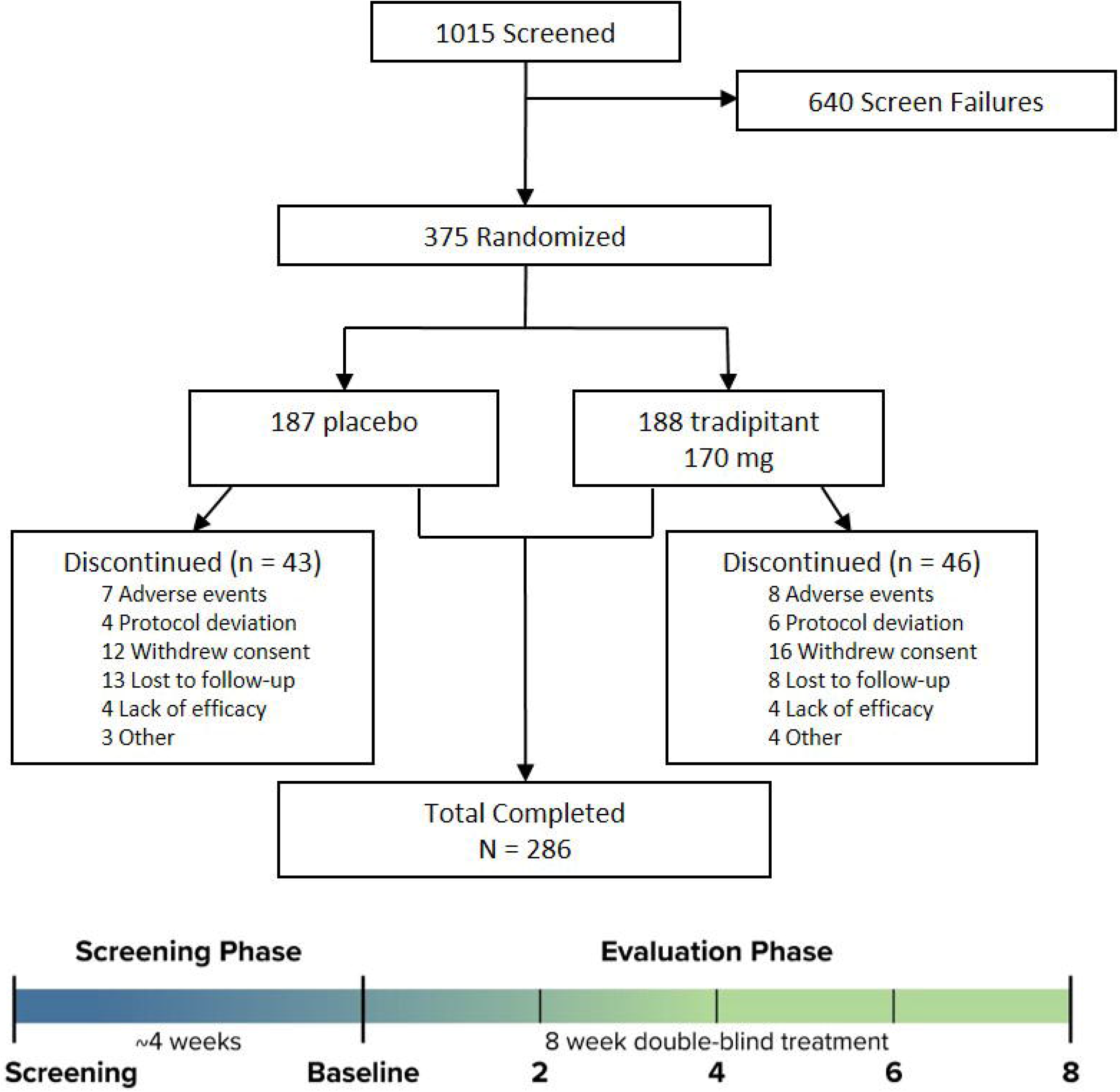
EPIONE Profile and Design.

Although there was a numerical benefit in the tradipitant group over placebo, EPIONE did not meet the primary endpoint of reduction in pruritus across the overall study population. Specifically, at Week 8, tradipitant and placebo patients both demonstrated significant and meaningful improvement in pruritus from baseline, but while the tradipitant magnitude of improvement was greater than that of placebo, the difference between treatment groups was not statistically significant (**Table II**, −0.2 (−0.8 to 0.4), P = 0.567). A significant interaction was observed between baseline disease severity (as defined by vIGA-AD score 0-4) and treatment (P < 0.001). This suggests that study participants with different baseline disease severities experienced different treatment outcomes. When adjusting for baseline disease severity and treatment in the ITT population, a significantly larger improvement in WI-NRS was seen in tradipitant-treated patients at Week 8 (**Table II**, −1.1 (−2.0 to −0.2), P = 0.022).

**Table II.** Efficacy Measures at Week 8. ^1^P values are from MMRM analysis. ^^^ITT n = 341, VLY-686 n = 171, PBO n = 170. ^#^IGA ≤ 2 n = 79, VLY-686 n = 40, PBO n = 39

| Endpoints<sup>1</sup> | Tradipitant | Placebo | Difference | P value |
| --- | --- | --- | --- | --- |
| **Itch Outcomes** |  |  |  |  |
| WI-NRS<sup>^</sup> | -3.61 | -3.43 | 0.18 | 0.5667 |
| WI-NRS Adjusting for IGA Severity<sup>^</sup> | -4.36 | -3.28 | 1.08 | 0.0217 |
| WI-NRS<sup>#</sup> | -4.74 | -3.14 | 1.60 | 0.0152 |
| Diary WI-NRS<sup>#</sup> | -4.23 | -2.14 | 2.09 | 0.0010 |
| **Disease Outcomes** |  |  |  |  |
| SCORAD Total<sup>#</sup> | -18.92 | -14.83 | -4.09 | 0.2466 |
| Objective SCORAD<sup>#</sup> | -9.94 | -9.22 | -0.72 | 0.7955 |
| Subjective SCORAD<sup>#</sup> | -8.81 | -5.79 | -3.02 | 0.0127 |
| EASI Total<sup>#</sup> | -2.19 | -1.62 | -0.57 | 0.3210 |
| **Sleep Outcomes** |  |  |  |  |
| SCORAD Sleep Disturbance<sup>#</sup> | -4.09 | -2.63 | -1.46 | 0.0131 |
| Diary Sleep Disturbance<sup>#</sup> | -3.12 | -1.98 | -1.14 | 0.0534 |
| POEM Sleep Disturbance<sup>#</sup> | -1.66 | -0.9 | -0.72 | 0.0086 |

A subgroup analysis showed that patients with mild disease severity (baseline vIGA-AD 1 or 2, n = 79; VLY-686 = 40, PBO = 39) had a significantly greater itch improvement when comparing tradipitant to placebo (−1.6 (−2.9 to −0.3), P = 0.015). Similar effects were seen throughout the treatment period at all post-randomization visits (Weeks 2, 4, 6, and 8) (**Figure 2 and 3a**). This treatment response in mild AD was confirmed by patient-reported daily diary worst itch improvement (**Table II**, −2.09 (−3.31 to −0.87), P = 0.001). Importantly, significant improvement in pruritus was observed after one day of tradipitant treatment (**Figure 3b**, −0.61 (−1.21 to −0.01), P = 0.0457). Two to four point improvement on the WI-NRS scale is considered clinically meaningful^32^. After eight weeks of treatment, 72.5% of tradipitant-treated patients achieved a reduction of 4 points or greater compared to 33.3% of placebo-treated patients (**Figure 3e**, 39.2 (18.9 to 59.4), P < 0.001).

**Figure 2.**
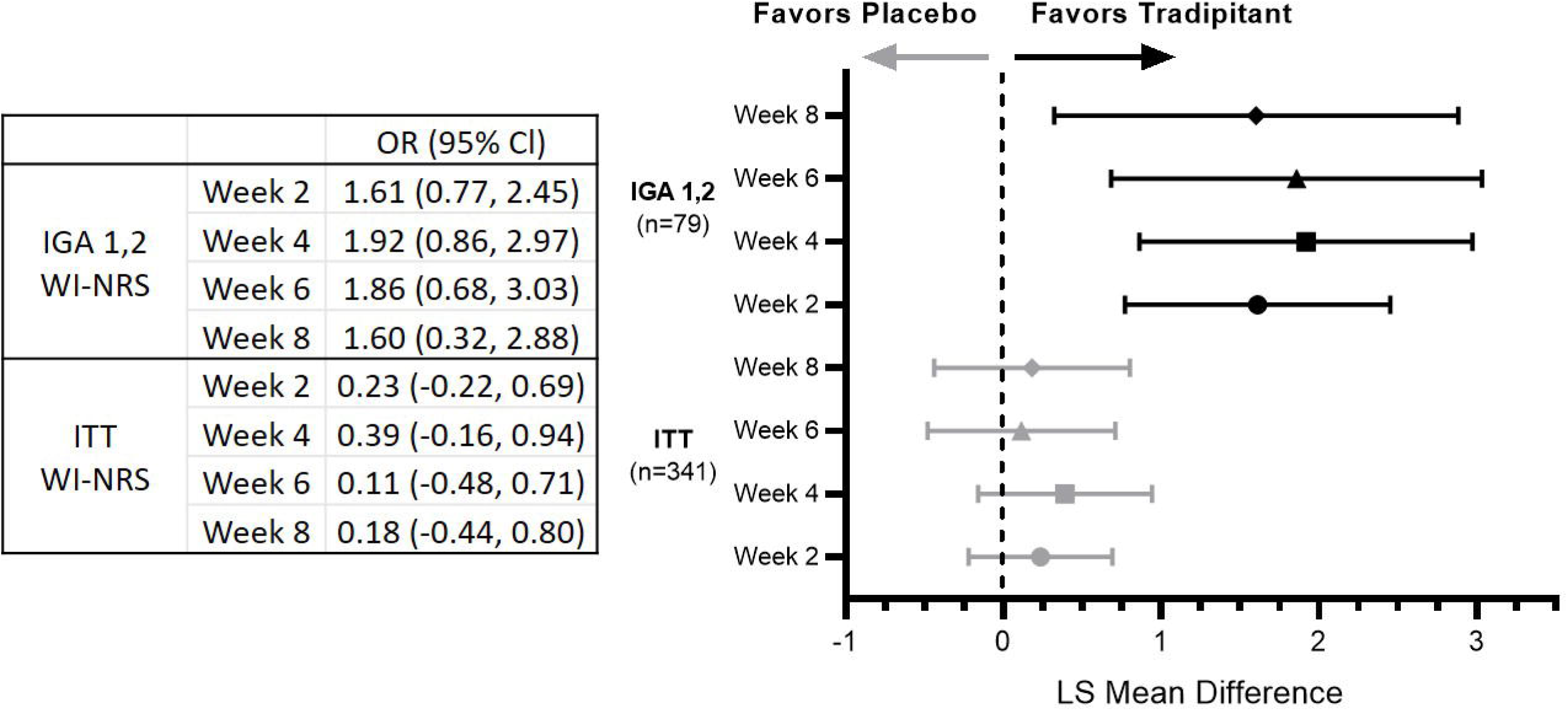
Worst Itch NRS Change by Week. Mild atopic dermatitis patients have greater improvement in worst itch after tradipitant-treatment. Forest plots of the analysis of ITT and IGA 1,2 WI-NRS change by week. Plotted as LS Mean Difference and 95% CI after tradipitant or placebo treatment.

**Figure 3.**
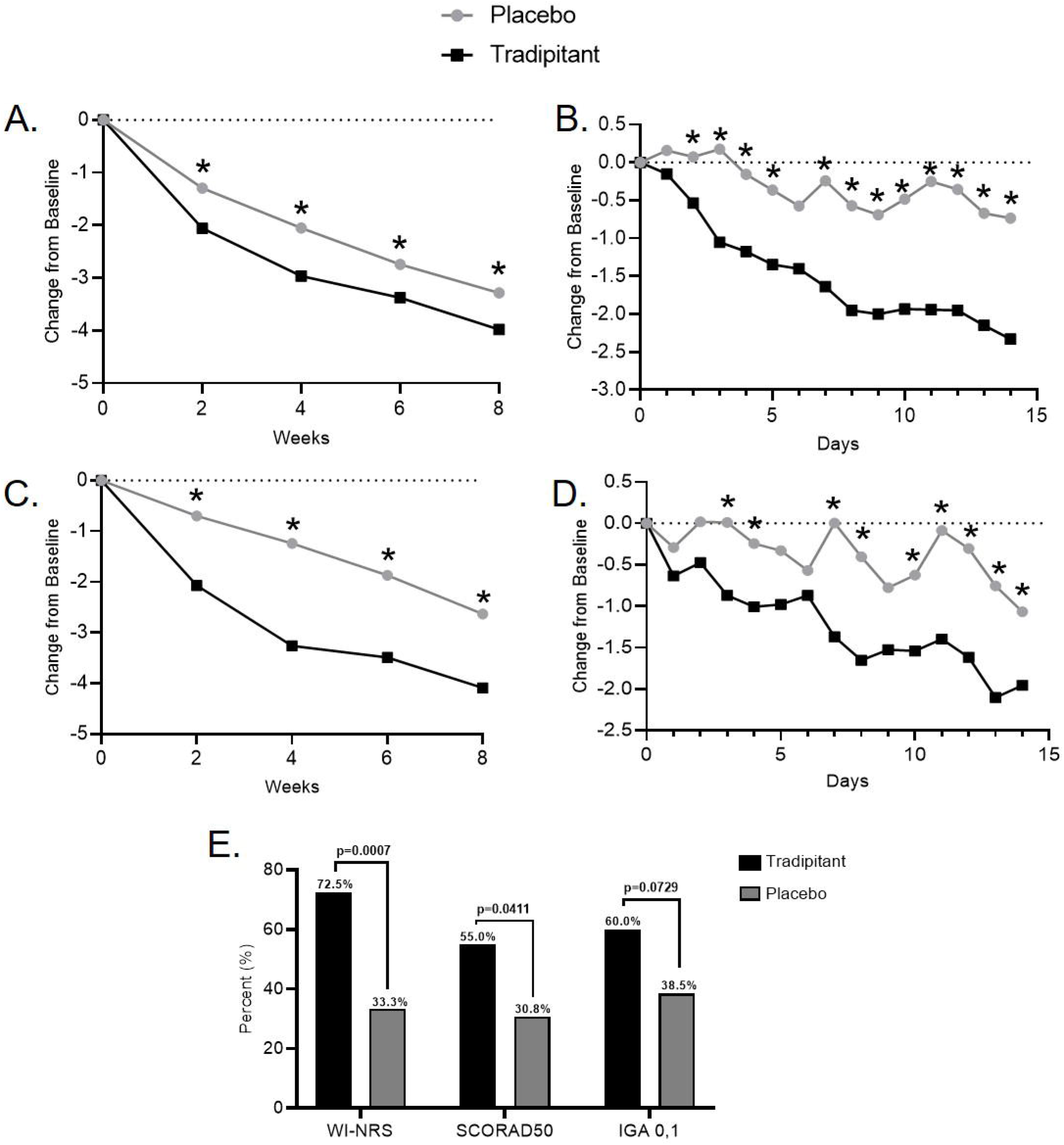
Tradipitant-treatment Improves Itch and Sleep in Mild Atopic Dermatitis. A. Tradipitant-treatment improved worst itch in mild AD. B. Improvement in itch was observed after one full day of tradipitant treatment. C. Tradipitant-treatment improved sleep disturbance in mild AD. D. Improvement in sleep was seen after two full days of tradipitant treatment. E. A greater proportion of mild AD patients achieved success of four points or greater on WI-NRS and at least a 50% improvement on SCORAD. A-D. P values are from MMRM analysis. E. P values are from Fisher’s exact test. *P < 0.05

Consistent with improvement on WI-NRS, mild AD patients also showed statistically significant improvement on the subjective SCORAD subscale relative to placebo (**Table II**, −3.02 (−5.37 to −0.66), P = 0.013). However, overall disease severity improvement was not seen in mild AD patients as measured by SCORAD (−4.09 (−11.07 to 2.89), P = 0.247) and EASI (−0.57 (−1.70 to 0.57), P = 0.321). In a responder analysis, 55% of tradipitant-treated patients saw their SCORAD reduced by at least half throughout the 8 weeks of treatment compared to 30.8% for those on placebo (**Figure 3e**, 24.2 (3.1 to 45.4), P = 0.041).

In addition to robust improvement in itch, improvement in nighttime sleep was also observed in mild AD. Mild AD patients experienced significant improvement in sleep as measured by the SCORAD VAS for average sleep disturbance over the last 72 hours after 8 weeks of tradipitant treatment (**Table II**, −1.46 (−2.60 to −0.32) P = 0.013). This improvement was confirmed by daily diary Sleep NRS (−1.14 (−2.3, 0.02), P = 0.053) and POEM sleep disturbance (−0.7 (−1.3 to −0.2), P = 0.009). A similar pattern was observed in SCORAD throughout the study (**Figure 3c**, Week 2 [-1.37 (−2.14, −0.61), P < 0.001] Week 4 [-2.01 (−3.04, - 0.99) P < 0.001]; Week 6 [-1.62 (−2.72, −0.53), P = 0.004]). Statistically significant improvement in sleep was seen after two full days of tradipitant treatment (**Figure 3d**, −0.88 (−1.58, −0.18), P = 0.0147).

The most frequent treatment emergent adverse events (TEAEs) were mild to moderate. There were no common TEAEs identified in the treatment arm as defined by > 5% incidence. TEAEs with > 2% incidence and twice that of placebo included diarrhea, fatigue, and worsening of AD. Severe TEAEs were reported in 5 patients, but were all determined to be unrelated to the study drug. No deaths were reported in the study.

## Discussion

In EPIONE, daily treatment of tradipitant for 8 weeks did not meet the primary endpoint of reduction in pruritus across the overall study population of patients with chronic pruritus related to AD, which included mild (23%), moderate (64%), and severe (13%) AD. However, tradipitant treatment resulted in a clinically meaningful reduction in patient reported worst itch and sleep disturbance in the mild AD study population. Statistically significant improvement was seen after a single day of treatment for itch and two days of treatment for sleep. Mild AD represents over 60% of the total AD population in the US^2^, thus this study potentially addresses a highly unmet need of treating pruritus for a large portion of AD patients that suffer from significant pruritus and sleep disturbance despite their mild lesions.

It is possible that the immediate and robust improvements observed in the mild AD subgroup were seen not only because of different levels of cutaneous inflammation but also because of distinct AD endotypes that are currently being defined for AD based on age, race, inflammatory, and genetic profiles^14,15^. We can confirm this hypothesis as Smieszek et al. (2020) showed that EPIONE patients with baseline IGA of 1,2 (mild) versus 3,4 (moderate to severe) have different sets of causative factors and courses, including different clinical manifestations, molecular levels, and genetic associations. The concept of different endotypes in AD underlines the need for targeted therapeutics in different AD disease types^14,15^.

Additionally, it is known that AD skin is enriched with hypersensitive sensory nerves that secrete elevated levels of SP^25^. These elevated levels of SP signal transmission of itch from peripheral nerves through the dorsal root ganglion to the brain where itch is perceived^8^. However, there are plenty of other itch mediators in AD including NGF, IL-2, IL-4, IL-13, and IL-31^8^. It is possible that in mild AD neuronal hypersensitivity and neuronal factors such as SP are the predominant pruriceptive mechanisms, while in moderate to severe AD immune mechanisms including interleukin release predominate and/or dilute the importance of SP signaling. As more pruritic factors propagate itch, the less efficacious a targeted therapy to reduce SP signaling may become. This may explain some of the effect observed in the overall study population in EPIONE.

Itch in AD is often worse at night^33^. Disturbed sleep has been reported in 33-81% of AD patients and is a major factor leading to impaired quality of life, and thus is a high unmet medical need^34^. After tradipitant treatment, improvement in sleep in mild AD patients was observed across three patient reported assessments, objective SCORAD VAS, daily diary, and POEM. Statistically significant improvement was seen after two days of treatment. Further study is needed to understand the minimal clinically important difference for these assessments to determine overall impact on quality of life. However, these results suggest that tradipitant can improve quality of life through the reduction of sleep disturbance.

Tradipitant may represent a new oral systemic option for mild AD patients based on the well-tolerated safety profile and immediate robust improvement in itch and sleep. Future studies are needed to confirm these efficacy results and refine treatment recommendations for AD patients who despite having mild lesions experience significant pruritus.

## Data Availability

Data available upon request

## Acknowledgements

We thank the investigators, study staff, and study patients who participated in this study.

## Abbreviations

AD: Atopic Dermatitis
AE: Adverse Event
BMI: Body Mass Index
BSA: Body Surface Area
EASI: Eczema Area and Severity Index
IGA: Investigator Global Assessment
ITT: Intent-to-Treat
LS: Mean Least Squared Mean
MMRM: Mixed Model Repeated Measures
NGF: Nerve Growth Factor
NK-1: Neurokinin-1
NRS: Numerical Rating Scale
POEM: Patient-Oriented Eczema Measure
SAE: Serious Adverse Event
SCORAD: SCORing Atopic Dermatitis
SP: Substance P
TEAE: Treatment Emergent Adverse Event
VAS: Visual Analogue Scale
vIGA-AD: Validated Investigator Global Assessment for Atopic Dermatitis
WI-NRS: Worst Itch Numerical Rating Scale

## References

1. Barbarot S, Auziere S, Gadkari A, et al. Epidemiology of atopic dermatitis in adults: Results from an international survey. Allergy Eur J Allergy Clin Immunol. 2018;(January):1–10. doi:10.1111/all.13401

2. Chiesa Fuxench ZC, Block J, Boguniewicz M, et al. Atopic Dermatitis in America Study: a cross-sectional study examining the prevalence and disease burden of atopic dermatitis in the US adult population. 2018. doi:10.1016/j.jid.2018.08.028

3. Correale CE, Walker C, Murphy L, Craig TJ. Atopic dermatitis: a review of diagnosis and treatment. AmFamPhysician. 1999;60(0002-838X (Print)):1110–1191.

4. Simpson EL, Bieber T, Eckert L, et al. Patient burden of moderate to severe atopic dermatitis (AD): Insights from a phase 2b clinical trial of dupilumab in adults. J Am Acad Dermatol. 2016;74(3):491–498. doi:10.1016/j.jaad.2015.10.043

5. Eichenfield LF, Tom WL, Chamlin SL, et al. Guidelines of care for the management of atopic dermatitis: Section 1. Diagnosis and assessment of atopic dermatitis. J Am Acad Dermatol. 2014;70(2):338–351. doi:10.1016/J.JAAD.2013.10.010

6. Huet F, Faffa MS, Poizeau F, Merhand S, Misery L, Brenaut E. Characteristics of pruritus in relation to self-assessed severity of atopic dermatitis. Acta Derm Venereol. 2019;99(3):279–283. doi:10.2340/00015555-3053

7. Dawn A, Papoiu ADP, Chan YH, Rapp SR, Rassette N, Yosipovitch G. Itch characteristics in atopic dermatitis: results of a web-based questionnaire. Br J Dermatol. 2009;160(3):642–644. doi:10.1111/j.1365-2133.2008.08941.x

8. Mollanazar NK, Smith PK, Yosipovitch G. Mediators of Chronic Pruritus in Atopic Dermatitis: Getting the Itch Out? Clin Rev Allergy Immunol. 2016;51(3):263–292. doi:10.1007/s12016-015-8488-5

9. Simpson EL, Guttman-Yassky E, Margolis DJ, et al. Association of inadequately controlled disease and disease severity with patient-reported disease burden in adults with atopic dermatitis. JAMA Dermatology. 2018;154(8):903–912. doi:10.1001/jamadermatol.2018.1572

10. Cabanillas B, Brehler A-C, Novak N. Atopic dermatitis phenotypes and the need for personalized medicine. Curr Opin Allergy Clin Immunol. 2017;17(4):309–315. doi:10.1097/ACI.0000000000000376

11. Eyerich K, Novak N. Immunology of atopic eczema: overcoming the Th1/Th2 paradigm. Allergy. 2013;68(8):974–982. doi:10.1111/all.12184

12. Sandilands A, Sutherland C, Irvine AD, McLean WHI. Filaggrin in the frontline: role in skin barrier function and disease. J Cell Sci. 2009;122(Pt 9):1285–1294. doi:10.1242/jcs.033969

13. O’Regan GM, Sandilands A, McLean WHI, Irvine AD. Filaggrin in atopic dermatitis. J Allergy Clin Immunol. 2009;124(3 Suppl 2):R2–6. doi:10.1016/j.jaci.2009.07.013

14. Czarnowicki T, He H, Krueger JG, Guttman-Yassky E. Atopic dermatitis endotypes and implications for targeted therapeutics. J Allergy Clin Immunol. 2019;143(1):1–11. doi:10.1016/j.jaci.2018.10.032

15. Brown SJ, McLean WHI. Eczema genetics: Current state of knowledge and future goals. J Invest Dermatol. 2009;129(3):543–552. doi:10.1038/jid.2008.413

16. Yosipovitch G, Papoiu ADP. What causes itch in atopic dermatitis? Curr Allergy Asthma Rep. 2008;8(4):306–311. doi:10.1007/s11882-008-0049-z

17. Wollenberg A, Oranje A, Deleuran M, et al. ETFAD/EADV Eczema task force 2015 position paper on diagnosis and treatment of atopic dermatitis in adult and paediatric patients. J Eur Acad Dermatology Venereol. 2016;30(5):729–747. doi:10.1111/jdv.13599

18. Wei W, Ghorayeb E, Andria ML, et al. 204 A real-world study evaluating adeQUacy of Existing Systemic Treatments for patients with moderate-to-severe Atopic Dermatitis (AD-QUEST): Baseline treatment patterns and unmet needs assessment. J Invest Dermatol. 2017;137(5):S35. doi:10.1016/j.jid.2017.02.219

19. Schmitt J, Csötönyi F, Bauer A, Meurer M. Determinants of treatment goals and satisfaction of patients with atopic eczema. JDDG. 2008;6(6):458–465. doi:10.1111/j.1610-0387.2007.06609.x

20. Teresiak-Mikołajczak E, Czarnecka-Operacz M, Jenerowicz D, Silny W. Neurogenic markers of the inflammatory process in atopic dermatitis: Relation to the severity and pruritus. Postep Dermatologii i Alergol. 2013;30(5):286–292. doi:10.5114/pdia.2013.38357

21. Nattkemper LA, Tey HL, Valdes-Rodriguez R, et al. The Genetics of Chronic Itch: Gene Expression in the Skin of Patients with Atopic Dermatitis and Psoriasis with Severe Itch. J Invest Dermatol. 2018;138(6):1311–1317. doi:10.1016/j.jid.2017.12.029

22. Almeida TA, Rojo J, Nieto PM, et al. Tachykinins and tachykinin receptors: structure and activity relationships. CurrMedChem. 2004;11(0929-8673 (Print)):2045–2081.

23. Johnson MB, Young AD, Marriott I. The Therapeutic Potential of Targeting Substance P/NK-1R Interactions in Inflammatory CNS Disorders. Front Cell Neurosci. 2017;10(January):1–14. doi:10.3389/fncel.2016.00296

24. Douglas SD, Leeman SE. Neurokinin-1 receptor: functional significance in the immune system in reference to selected infections and inflammation. Ann N Y Acad Sci. 2011;1217:83–95. doi:10.1111/j.1749-6632.2010.05826.x

25. Zhang Z, Zheng W, Xie H, et al. Up-regulated expression of substance P in CD8+ T cells and NK1R on monocytes of atopic dermatitis. J Transl Med. 2017;15(1):93. doi:10.1186/s12967-017-1196-6

26. Wei T, Guo T-Z, Li W-W, Hou S, Kingery WS, Clark JD. Keratinocyte expression of inflammatory mediators plays a crucial role in substance P-induced acute and chronic pain. J Neuroinflammation. 2012;9:181. doi:10.1186/1742-2094-9-181

27. Ohmura T, Hayashi T, Satoh Y, Konomi A, Jung B, Satoh H. Involvement of substance P in scratching behaviour in an atopic dermatitis model. Eur J Pharmacol. 2004;491(2-3):191–194. doi:10.1016/j.ejphar.2004.03.047

28. Heitman A, Xiao C, Cho Y, Polymeropoulos C, Birznieks G, Polymeropoulos MH. Tradipitant improves worst itch and disease severity in patients with chronic pruritus related to atopic dermatitis. J Am Acad Dermatol. 2018;79(3):AB300. doi:10.1016/j.jaad.2018.05.1184

29. Oranje AP, Glazenburg EJ, Wolkerstorfer A, de Waard-van der Spek FB. Practical issues on interpretation of scoring atopic dermatitis: the SCORAD index, objective SCORAD and the three-item severity score. Br J Dermatol. 2007;157(4):645–648. doi:10.1111/j.1365-2133.2007.08112.x

30. Hanifin JM, Thurston M, Omoto M, Cherill R, Tofte SJ, Graeber M. The eczema area and severity index (EASI): assessment of reliability in atopic dermatitis. EASI Evaluator Group. ExpDermatol. 2001;10(0906-6705 (Print)):11–18.

31. Charman CR, Venn AJ, Williams HC. The patient-oriented eczema measure: development and initial validation of a new tool for measuring atopic eczema severity from the patients’ perspective. Arch Dermatol. 2004;140(12):1513–1519. doi:10.1001/archderm.140.12.1513

32. Yosipovitch G, Reaney M, Mastey V, et al. Peak Pruritus Numerical Rating Scale: psychometric validation and responder definition for assessing itch in moderate-to-severe atopic dermatitis. Br J Dermatol. 2019;181(4):761–769. doi:10.1111/bjd.17744

33. Chang Y Sen, Chiang BL. Mechanism of sleep disturbance in children with atopic dermatitis and the role of the circadian rhythm and melatonin. Int J Mol Sci. 2016;17(4). doi:10.3390/ijms17040462

34. Jeon C, Yan D, Nakamura M, et al. Frequency and Management of Sleep Disturbance in Adults with Atopic Dermatitis: A Systematic Review. Dermatol Ther (Heidelb). 2017;7(3):349–364. doi:10.1007/s13555-017-0192-3

